# Relationships between 25-Hydroxyvitamin D and Alpha-synucleinopathies: A Mendelian Randomization Study

**DOI:** 10.1101/2024.11.25.24317894

**Authors:** Fu-Jia Li, Ru-Lin Geng, Cheng Wen, Kai-Xun Huang, Dan-Yu Lin, Zhong-Gui Li, Jie-Li Zhang, Chuan-Ying Xu, Wei Zhang, Jie Zu, Li-Guo Dong, Wen-Xin Wang, Zhi-Jia Ruan, Pei-Xiao Yin, Chen-Yang Guan, Gui-Yun Cui, En-Xiang Tao

## Abstract

**Background:** Alpha-synucleinopathies, including Parkinson’s disease (PD), multiple system atrophy (MSA), and dementia with Lewy bodies (DLB), are neurodegenerative diseases characterized by abnormal aggregation of alpha-synuclein. Isolated REM sleep behavior disorder (iRBD) is a prodromal stage of these disorders. While 25-hydroxyvitamin D (25[OH]D) has been linked to PD in previous studies, its causal role remains unclear, and its relationship with iRBD, MSA, and DLB is rarely explored.

**Objectives:** This study used Mendelian randomization (MR) to investigate the causal relationship between 25(OH)D and iRBD, PD, MSA, and DLB.

**Methods:** GWAS data for 25(OH)D were sourced from the UK Biobank, and outcome data for PD were obtained from the latest FinnGen R12 database. Data for iRBD, PD onset age, PD clinical progression, MSA, and DLB were derived from a broader European population. The study had two steps: univariable MR (UVMR) using genome-wide IV selection as the primary analysis, with replication analyses using biological function-based IV selection and sunlight exposure and vitamin D supplementation-related factors as exposures. Multivariable MR (MVMR) was then conducted to adjust for 13 covariates, testing the robustness of findings.

**Results:** 25(OH)D showed a significant protective effect on MSA across all UVMR (both primary and replication) and MVMR analyses. It also exhibited a nominal protective effect on PD in the Finnish population in primary UVMR analysis, but this was not replicated. No associations were found between 25(OH)D and iRBD, PD onset age, PD clinical progression, or DLB. Power analyses indicated insufficient sensitivity to detect weak effects of 25(OH)D on iRBD and DLB.

**Conclusions:** MR analysis supports a protective role of 25(OH)D against MSA. While nominally protective for PD in the Finnish population, the effect was not robust. Weak effects on iRBD and DLB cannot be excluded.

## Introduction

Alpha-synucleinopathies, encompassing Parkinson’s disease (PD), multiple system atrophy (MSA), and dementia with Lewy bodies (DLB), are neurodegenerative disorders characterized by the misfolding and abnormal aggregation of alpha-synuclein^1^. These diseases result in progressive disability and mortality, with overlapping yet distinct clinical manifestations. PD presents with characteristic motor symptoms such as bradykinesia, resting tremor, and rigidity^2^. MSA is defined by severe autonomic dysfunction, often accompanied by parkinsonism or ataxia^3^. In contrast, DLB is marked by fluctuating cognitive decline, visual hallucinations, and parkinsonism^4^. Notably, isolated rapid eye movement sleep behavior disorder (iRBD), a condition characterized by the loss of muscle atonia during REM sleep and the presence of abnormal behaviors^5^, is recognized as a prodromal phase of alpha-synucleinopathies^6^. Despite the substantial burden on patients and healthcare systems, effective therapies for alpha-synucleinopathies remain scarce. Therefore, identifying modifiable risk factors is critical for prevention and improving patient outcomes.

Vitamin D, a fat-soluble steroid hormone, primarily exists in the body as 25-hydroxyvitamin D (25[OH]D). It has been suggested to protect against neurodegenerative diseases by maintaining calcium homeostasis, reducing oxidative stress, inhibiting inflammation, and preventing the aggregation of misfolded proteins (including alpha-synuclein) ^7,8^, which are both involved in the pathogenesis of alpha-synucleinopathies. However, whether 25(OH)D provides clinical protection against alpha-synucleinopathies remains unclear.

In PD, most cross-sectional studies report lower 25(OH)D levels in patients compared to healthy controls^9–11^, but it is debated whether this reduction contributes to disease onset or results from disease progression. Immobility in PD could limit sunlight exposure, as indicated by reduced outdoor time, thereby decreasing vitamin D synthesis. Additionally, gastrointestinal dysfunction may impair absorption, both of which could potentially lead to lower 25(OH)D levels. Although cohort studies reduce reverse causation bias, their findings are inconsistent^12–14^. Additionally, clinical trials on vitamin D supplementation have not consistently shown PD symptom improvement^15–17^. Research on vitamin D in MSA^18,19^ and DLB^20^ is even more limited, hindered by small sample sizes and methodological issues, such as cross-sectional designs and inadequate control for confounders like outdoor time and dietary vitamin D intake. These limitations challenge the reliability and generalizability of existing findings.

To develop effective interventions, identifying risk factors within the causal pathway of the disease is essential. Mendelian randomization (MR) provides a robust method for causal inference by minimizing confounding bias and reverse causation. The limitations of the existing MR study, however, highlight the need for further research. In 2017, Susanna C. Larsson and colleagues used MR to examine the relationship between 25(OH)D and PD but found no causal link^21^. They partly attributed the inconclusive results to the small effect size of 25(OH)D and the limited number of instrumental variables (IVs), which reduced statistical power.

With the availability of large genome-wide association study (GWAS) datasets and advancements in MR methods, this study aims to comprehensively investigate the relationships between 25(OH)D levels and iRBD, PD, age at PD onset, PD clinical progression, MSA, and DLB.

## Method

### 1. Study Design

Our study consisted of two steps. Step one involved univariable MR (UVMR) analysis, where we selected IVs across the whole genome to investigate the effects of 25(OH)D on the following outcomes: iRBD, PD, age at PD onset, progression of 21 clinical PD phenotypes, MSA, and DLB. This served as the primary analysis.

Outcomes showing nominal statistical significance with 25(OH)D (P < 0.05) underwent further replication analysis, with biological function-based IV selection and sunlight exposure and vitamin D supplementation-related factors as exposures. In step two, outcomes with consistent results in primary and replication UVMR analyses were further included in the MVMR analysis to adjust for covariates. The study strictly adhered to the three core assumptions^22^ of MR analysis and followed the STROBE-MR guidelines^23^ (**Table S2**).

### 2. Data Source

We obtained summary-level GWAS statistics from recent large-scale studies of European populations. To minimize sample overlap, exposure and outcome data were derived from different cohorts. Ethical approvals were obtained from each participating cohort, so no additional ethical approvals or informed consents were required. The composition of GWAS data cohorts, the content of questionnaires used to define clinical characteristics, the specific methods and kits used for laboratory measurements, and the diagnostic criteria for diseases can be found in **Table S1**.

#### 2.1 Exposures

As in previous studies^24–26^, we used the most recent 25(OH)D GWAS dataset, which is based on the UK Biobank population (n = 417,580)^27^. Serum 25(OH)D concentrations were measured using a chemiluminescence immunoassay (Diasorin Liaison®), which covered 25(OH)D3 and 25(OH)D2. Individuals with total concentrations outside the range of 10–375 nmol/L were excluded. The median, mean, and interquartile range of 25(OH)D concentrations were 47.9, 49.6, and 33.5–63.2 nmol/L, respectively.

Most vitamin D is synthesized in the skin through UV exposure, with only ∼20% obtained from dietary intake^28^. Similar to previous observational studies^11,29^, our study also considered characteristics related to sunlight exposure and vitamin D supplementation as exposure factors. The GWAS data for these characteristics were derived from the UK Biobank and included variables such as time spend outdoors in summer (n = 419,314) and winter (n = 364,465), skin color (n = 456,692), use of sun protection (n = 459,416), use of vitamin D supplements (cases/controls = 17,879/442,472), and non-use of any vitamin or mineral supplements (cases/controls = 144,656/315,695). These data were self-reported through touchscreen questionnaires.

#### 2.2 Covariates

The “*2024 Consensus Statement on Vitamin D Status Assessment and Supplementation*”^30^ highlighted key factors influencing vitamin D levels. If genetic IVs for 25(OH)D are strongly associated with these factors, they may directly impact outcomes via non-25(OH)D pathways, introducing horizontal pleiotropy and biasing the results. To mitigate this, we identified 13 covariates, grouped them into three sets, and sequentially incorporated them into MVMR analyses to minimize horizontal pleiotropy and strengthen the reliability of our causal inference. The covariate data were mainly derived from the UK Biobank. Set 1 included lifestyle and socioeconomic factors: body mass index, smoking initiation, alcohol intake frequency, levels of moderate-to-vigorous physical activity, and years of education. Set 2 comprised vitamin D metabolism-related markers: alanine aminotransferase, creatinine, and parathyroid hormone levels. Set 3 included chronic health conditions: hypertension, type 2 diabetes, atherosclerotic cardiovascular diseases, stroke, and gastrointestinal or abdominal diseases.

#### 2.3 Outcomes

To avoid sample overlap, given that the exposure data in this study were sourced from the UK Biobank, we did not use datasets curated by Mike A. Nalls (December 2019)^31^ or Jonggeol Jeffrey Kim (January 2024)^32^. Instead, we used the latest PD GWAS data from the FinnGen R12 database (https://r12.finngen.fi/, November 2024), which included 5,861 cases and 494,487 controls. PD diagnoses were based on ICD-10 (G20), ICD-9 (3320A), and ICD-8 (34200) codes. Similarly, Tao Wang et al. followed the same data selection strategy under similar circumstances^33^. The PD age-at-onset dataset was obtained from the International Parkinson’s Disease Genomics Consortium^34^. Age at onset was defined based on patient-reported initial parkinsonian motor symptoms, or, if unavailable, the age of diagnosis was used as a proxy. The datasets on PD clinical progression included 12 longitudinal cohorts of PD patients recruited from North America, Europe, and Australia^35^. These datasets included 21 clinical features of disease progression, covering changes in staging, worsening motor symptoms, reduced daily functioning, aggravated non-motor symptoms, cognitive decline, new motor complications, depression, impaired smell, increased constipation, and sleep disturbances. The iRBD GWAS data, aggregated by Lynne Krohn’s team in December 2022, included 1,061 cases and 8,386 controls from cohorts in France, Canada, Italy, the UK, and other European countries^36^. Cases were identified by the International RBD Study Group, diagnosed according to the International Classification of Sleep Disorders (2nd or 3rd Edition), and confirmed via video-polysomnography. MSA GWAS data, aggregated by Ruth Chia’s team in July 2024, included 888 cases and 7,128 controls recruited from 20 centers across North America and Europe^37^. Cases were diagnosed as clinically probable or pathologically confirmed using the Gilman consensus criteria. DLB GWAS data, also aggregated by Ruth Chia’s team in February 2021, comprised 2,591 cases and 4,027 controls^38^. Participants were recruited from 44 centers in Europe and North America, with controls drawn from the Wellderly cohort and the National Institute on Aging. DLB cases were either autopsy-confirmed or clinically probable.

### 3. Genetic Instrument Selection

In this study, IVs were selected using two approaches. First, genome-wide IV selection: single nucleotide polymorphisms (SNPs) significantly associated with the exposure (P < 5 × 10□□) across the whole genome were chosen. If this threshold yielded insufficient IVs, a more lenient threshold (P < 5 × 10□□) was applied, selecting IVs for iRBD, PD age at onset, progression of PD symptoms, MSA, DLB, and vitamin D supplementation. Second, biological function-based IV selection: IVs were selected from genes involved in vitamin D metabolism, including GC, CYP2R1, DHCR7, and CYP24A1^39,40^.

The R² value was used to assess the proportion of variance in exposure explained by each SNP, while the F-statistic ensured instrument strength (retaining only SNPs with F > 10). SNPs in linkage disequilibrium (r² > 0.001, window size 10,000 kb) or with effect allele frequencies below 0.01 were excluded. To meet MR independence assumptions, SNPs directly associated with the outcome were identified and excluded according to the LDtrait database (https://ldlink.nih.gov/?tab=ldtrait).

### 4. Mendelian Randomization

For the UVMR analyses, we used multiple analytical models. The inverse variance weighted (IVW)^41^ method served as the primary model due to its high statistical power when all instrumental variables are valid, though it is sensitive to heterogeneity and pleiotropy. To enhance robustness, we applied five secondary models: MR-Egger^42^ and weighted median^43^ methods, which provide reliable estimates in the presence of pleiotropy or invalid instruments, albeit with lower power; the constrained maximum likelihood-model average (cML-MA)^44^ method, robust against pleiotropy; the robust adjusted profile score (MR-RAPS)^45^ method, effective for weak instruments; and Bayesian weighted MR (BWMR)^46^, which uses Bayesian weighting to address pleiotropy and accounts for polygenicity, making it suitable for analyzing complex traits or diseases. These methods complemented each other and contributed to the reliability and robustness of the results. MR results were reported as odds ratios (OR) with 95% confidence intervals (CI), representing the effect of a genetically determined unit change in exposure on outcome risk. For continuous exposures, the OR reflects the change in risk per one standard deviation increase; for binary exposures, it represents the risk ratio between exposed and non-exposed groups.

In the UVMR analyses, several sensitivity analyses were performed to ensure robustness: (1) Results from genome-wide IV selection served as the primary UVMR analysis, while replication analyses used biological function-based IV selection and exposures such as sunlight exposure and vitamin D supplementation for mutual validation; (2) Effect estimates were obtained using one primary model and five secondary models suited to different assumptions; (3) Cochran’s Q-statistic was applied to evaluate heterogeneity in the IVW results; (4) Potential horizontal pleiotropy was assessed using the MR-Egger intercept p-value^42^ and the Mendelian Randomization Pleiotropy RESidual Sum and Outlier (MR-PRESSO) global test^47^; (5) Outliers among IVs were identified using MR-PRESSO and MR-Radial (Radial-IVW and Radial-Egger tests, with P < 0.05 indicating outliers), and MR analyses were repeated after removing identified outliers; (6) Leave-one-out analyses were conducted to determine whether the results were driven by specific SNPs; (7) Finally, the tool https://shiny.cnsgenomics.com/mRnd/ was used to calculate the minimum detectable causal effect size at 80% statistical power in the MR analyses^48^.

To reduce bias from reverse causality, we adopted the following strategies: (1) Excluding IVs significantly associated with the outcome; (2) Testing the causal direction using the Steiger test^49^; and (3) Conducting reverse MR analyses.

In the MVMR analyses, causal effects were estimated using the MVMR-IVW, and MVMR-Egger methods, with horizontal pleiotropy evaluated based on whether the MR-Egger intercept significantly deviated from zero. When no pleiotropy was detected, the MVMR-IVW method was used as the primary estimator; if pleiotropy was present, the results primarily relied on the MVMR-Egger method^42^.

All analyses were two-sided tests performed using R version 4.4.1. Relevant R packages included TwoSampleMR, MendelianRandomization, MR-PRESSO, RadialMR, MR.RAPS, MRcML, and BWMR.

## Results

### 1. Primary UVMR analysis results

After quality control and removing SNPs in linkage disequilibrium, 115 SNPs strongly associated with 25(OH)D were retained as candidate IVs (**Table S3**). A search on the LDtrait platform identified six SNPs directly associated with alpha-synucleinopathies (**Table S4**), which were excluded. Among the remaining 109 SNPs, 104, 104, 97, 97, and 98 were successfully harmonized with the datasets for iRBD, PD, age at onset of PD, MSA, and DLB, respectively. These SNPs explained approximately 4.28%-4.42% of the variance in 25(OH)D levels, and all F-statistics exceeded 10, confirming that the MR results were not affected by weak instruments (**Table 1**).

UVMR analysis showed a significant protective effect of higher 25(OH)D levels on MSA (OR [95% CI]: 0.598 [0.421, 0.851], P = 0.004, P_FDR = 0.021, **Figure 2**). This finding was supported by the primary analysis model (IVW) and four secondary models (MR Egger, Weighted median, MR-RAPS, and BWMR), with the cML-MA model showing consistent effect direction despite not reaching statistical significance. MR-PRESSO identified no significant outliers among the 97 instrumental variables, while MR-Radial detected 1 potential outlier (**Table S5**). Reanalysis excluding this outlier produced robust results (**Table S6**). Steiger directionality tests and reverse MR (**Table S9, 10**) supported the causal direction, and Cochran’s Q test indicated no heterogeneity. MR-Egger regression and the MR-PRESSO global test showed no evidence of horizontal pleiotropy (**Table S7**). Leave-one-out analysis confirmed that results were not driven by individual SNPs (**Table S8**).

MR analysis also suggested a nominal protective effect of higher 25(OH)D levels on PD (OR [95% CI]: 0.867 [0.758, 0.991], P = 0.036, P_FDR = 0.091, **Figure 2**), supported by the IVW, MR-RAPS, and BWMR models, with consistent effect directions across all six models. MR-PRESSO detected no significant outliers among the 104 instrumental variables, while MR-Radial identified 7 potential outliers (**Table S5**). Reanalysis excluding these outliers showed robust results (**Table S6**). Sensitivity analyses confirmed the findings were consistent in direction and unaffected by heterogeneity or horizontal pleiotropy (**Table S7-10**).

However, UVMR analysis did not provide evidence of an effect of 25(OH)D levels on age at onset of PD, PD clinical progression (**Table S6**), or the risk of iRBD and DLB (**Figure 2**). These results remained robust in sensitivity analyses (**Table S7-10**). Statistical power calculations suggested that UVMR analysis might not have been sufficient to detect weaker effects of 25(OH)D on iRBD and DLB (**Table 1**).

### 2. Replication UVMR analysis results

We selected eight SNPs from four key genes involved in vitamin D metabolism as IVs (GC: rs111978466, rs1352846, rs62318873; DHCR7: rs2276360, rs36037728; CYP2R1: rs116970203; CYP24A1: rs8121940, rs8123293, **Table S11**) and conducted UVMR analysis again. The results confirmed that 25(OH)D remains a protective factor for MSA (OR [95% CI]: 0.562 [0.366, 0.863], P = 0.008, **Figure 3A**). However, its effect on PD was no longer significant. We also examined the relationships between sunlight exposure, vitamin D supplementation-related factors, and MSA or PD. Darker skin and “non-use of vitamin or mineral supplements” were identified as risk factors for MSA, while vitamin D supplementation had a protective effect on MSA (**Figure 3B**, **Table S12**). No evidence was found to support an influence of sunlight exposure or vitamin D supplementation on PD risk. These findings remained robust in sensitivity analyses (**Table S13**).

### 3. MVMR analysis results

After adjusting for lifestyle and socioeconomic factors (body mass index, smoking, alcohol intake frequency, moderate-to-vigorous physical activity levels, and years of education), vitamin D metabolism-related markers (alanine aminotransferase, creatinine, and parathyroid hormone levels), and chronic health conditions (hypertension, type 2 diabetes, atherosclerotic cardiovascular disease, stroke, and gastrointestinal or abdominal disease), the protective effects of 25(OH)D on MSA remained significant (**Figure 4**). Sensitivity analyses suggested that some results were mildly influenced by heterogeneity (**Table S14**), likely due to the large number of instrumental variables used. None of the results were affected by horizontal pleiotropy (**Table S14**).

## Discussion

This is the first MR study comprehensively exploring the relationship between serum 25(OH)D levels and alpha-synucleinopathies. Our findings provide evidence for the protective effect of 25(OH)D on MSA, which remained robust across primary and replication UVMR analyses and after adjusting for 13 covariates in MVMR analysis. Additionally, 25(OH)D showed a nominal protective effect on PD in the Finnish population, though this lacked sufficient robustness. The possibility of a weak effect on iRBD or DLB cannot be ruled out.

### 1. Declined 25(OH)D Levels in PD: Risk Factor or Consequence?

PD is the second most common neurodegenerative disorder and the most prevalent α-synucleinopathy. The relationship between 25(OH)D and PD remains controversial. Most cross-sectional studies have reported significantly lower 25(OH)D levels in PD patients compared to controls^9–11^. However, this decrease may reflect disease progression rather than causality, potentially influenced by reduced sunlight exposure due to impaired mobility or vitamin D malabsorption caused by gastrointestinal symptoms. In contrast, cohort studies are less affected by reverse causation but yield inconsistent findings. While studies in Finnish and UK Biobank cohorts support a protective role of 25(OH)D against PD^12,13^, research from the United States found no significant association^14^. Variability in genetic background, geographic factors, and behavioral differences across cohorts likely contributes to these discrepancies. MR analysis offers a robust approach to address confounding and reverse causation compared to observational studies. In 2017, Susanna C. Larsson utilized small GWAS data in an MR analysis of 25(OH)D and PD but found no significant association^21^. Using larger GWAS datasets, our study identified a nominal protective effect of 25(OH)D against PD in the Finnish population. However, this finding lacked robustness in replication analyses. We speculate that the protective effect of 25(OH)D on PD may be modest, and the limited statistical power of replication analyses might have hindered the detection of such a weak effect, which could also explain the null results in Larsson’s study.

Notably, a 2011 cohort study reported that 25(OH)D levels did not decline with PD progression^50^. Similarly, our reverse MR analysis showed that PD status does not significantly affect 25(OH)D levels. Furthermore, advancing Hoehn and Yahr stages, worsening motor symptoms, and increased severity of constipation were not associated with changes in 25(OH)D levels. These findings provide additional evidence supporting the hypothesis that reduced 25(OH)D levels are a risk factor for PD rather than a consequence of disease progression.

### 2. 25(OH)D and MSA: Present Evidence and Potential Mechanisms

MSA is at least ten times less prevalent than PD but progresses more rapidly, often causing severe disability within 5–6 years and death within 10 years of symptom onset^3^. Due to its rarity, only two cross-sectional studies have investigated the relationship between 25(OH)D levels and MSA, both reporting significantly lower levels in MSA patients compared to controls^18,19^. However, these studies did not adjust for confounders, limiting their reliability. Our MR analysis supports a causal relationship between lower 25(OH)D levels and increased MSA risk. This conclusion was robust across primary and replication UVMR analyses and remained consistent after comprehensive covariate adjustment using MVMR. The MR findings align with trends observed in previous studies but should be interpreted cautiously due to population differences. Observational studies were predominantly conducted in Asian cohorts, while our MR analysis used European data. Notably, genetic differences and subtype distributions (MSA-P vs. MSA-C) between European and Asian MSA patients^3^ may influence results. Prospective observational studies in European populations are needed to validate these findings.

Vitamin D can cross the blood-brain barrier^51^, and its receptors are widely expressed in neurons and glial cells^52^, providing a biological basis for its potential role in MSA. Research suggests that vitamin D exerts neuroprotective effects in neurodegenerative diseases by suppressing immune-inflammatory responses, reducing oxidative stress, enhancing autophagy, and promoting neurotrophic factor expression^7,53^. These mechanisms may also explain its protective role in MSA, which shares overlapping pathogenic pathways with other neurodegenerative disorders^3,54^. Additionally, 25(OH)D may also confer protective effects against MSA through distinct mechanisms. MSA is characterized by oligodendroglial cytoplasmic inclusions (GCIs) rich in aggregated α-synuclein, along with extensive myelin degeneration and oligodendrocyte dysfunction (impaired oligodendrocyte precursor cell [OPC] maturation, remyelination defects, and reduced neuroprotective functions)^54–58^. In vitro studies demonstrate that vitamin D reduces α-synuclein oligomer aggregation and associated neurocytotoxicity in neurons^8^, though its potential effect on α-synuclein deposition in oligodendrocytes remains unclear. Additionally, vitamin D promotes remyelination and neurorepair by enhancing OPC differentiation and creating a protective microenvironment^59–62^, suggesting its therapeutic potential in MSA.

### 3. Clinical Implications

(1) MSA progresses rapidly, with no disease-modifying treatments or effective symptomatic management. Reducing its incidence is essential to alleviating the disease burden. While risk factors for MSA were previously unknown, our study is the first to identify 25(OH)D as a protective factor. Given the high prevalence of vitamin D deficiency^63^, widespread vitamin D supplementation may have the potential to significantly reduce MSA incidence. (2) Our findings highlight the need for multicenter prospective studies to confirm the protective effect of 25(OH)D on MSA. Further exploration is warranted to determine if this effect extends to patients with iRBD and early autonomic dysfunction, prodromal stages of MSA^6^. (3) Our study provides additional evidence that reduced 25(OH)D levels are a risk factor for PD rather than a consequence of disease progression, requiring further follow-up studies.

### 4. Limitations

(1) Our study population was European, which may limit the generalizability of the findings. While we identified a protective effect of 25(OH)D against MSA, previous supporting observational studies were conducted in Asian populations. Future observational studies in European populations are needed to validate this conclusion. (2) The limited sample size prevented us from determining whether the lack of associations between 25(OH)D, iRBD risk, PD progression, and DLB risk reflects insufficient statistical power or a true absence of association. Larger GWAS datasets are needed for future MR analyses. (3) Due to data constraints, we could not assess whether 25(OH)D influences MSA risk in patients with iRBD or early autonomic dysfunction, nor its effects on MSA progression. These areas warrant further study.

### 5. Conclusions

In summary, our study identifies a protective role of 25(OH)D against MSA. Although 25(OH)D showed a nominal protective effect on PD in the Finnish population, the result lacked robustness. Potential weak effects on iRBD and DLB cannot be excluded.

## Data Availability

The GWAS data sources used in this study are detailed in the Methods section and Table S1. Code book, and analytic code will be made available upon request pending.

## Acknowledgement

FJL, RLG, and CW designed research; FJL, RLG, CW, GYC, EXT, and the other authors (KXH, DYL, ZGL, JLZ, CYX, WZ, JZ, LGD, WXW, ZJR, PXY, CYG) conducted research; GYC, EXT, and the other authors (KXH, DYL, ZGL, JLZ, CYX, WZ, JZ, LGD, WXW, ZJR, PXY, CYG) provided essential materials; FJL, RLG, and CW analyzed data; and FJL, RLG, and CW wrote the paper. GYC and EXT had primary responsibility for final content. All authors read and approved the final manuscript.

## Conflict of Interest

The authors have no conflicts of interest relevant to the content of this article.

## Funding

This study was funded by Shenzhen Science and Technology Program (JCYJ20220818102206014), National Natural Science Foundation of China (82271454), the Futian Healthcare Research Project (FTWS2022010), the District Key Specialty Funds / Department of Neurology (QZDZK-202410), and the Hospital Key Discipline Funds / Department of Neurology (YZDXKJF-202017).

## Abbreviations

25(OH)D: 25-hydroxyvitamin D
BWMR: Bayesian Weighted Mendelian randomization
CI: Confidence interval
cML-MA: Constrained maximum likelihood-model average
DLB: Dementia with Lewy bodies
GCI: Oligodendroglial cytoplasmic inclusion
IV: Instrumental variable
IVW: Inverse variance weighted
iRBD: Isolated rapid eye movement sleep behavior disorder
LD: Linkage disequilibrium
MR: Mendelian randomization
MR-PRESSO: Mendelian randomization pleiotropy residual sum and outlier
MR-RAPS: Mendelian randomization robust adjusted profile score
MSA: Multiple system atrophy
nSNPs: Number of instrumental variables
OPC: Impaired oligodendrocyte precursor cell
OR: Odds ratio
PD: Parkinson’s disease
SNP: Single nucleotide polymorphism

---

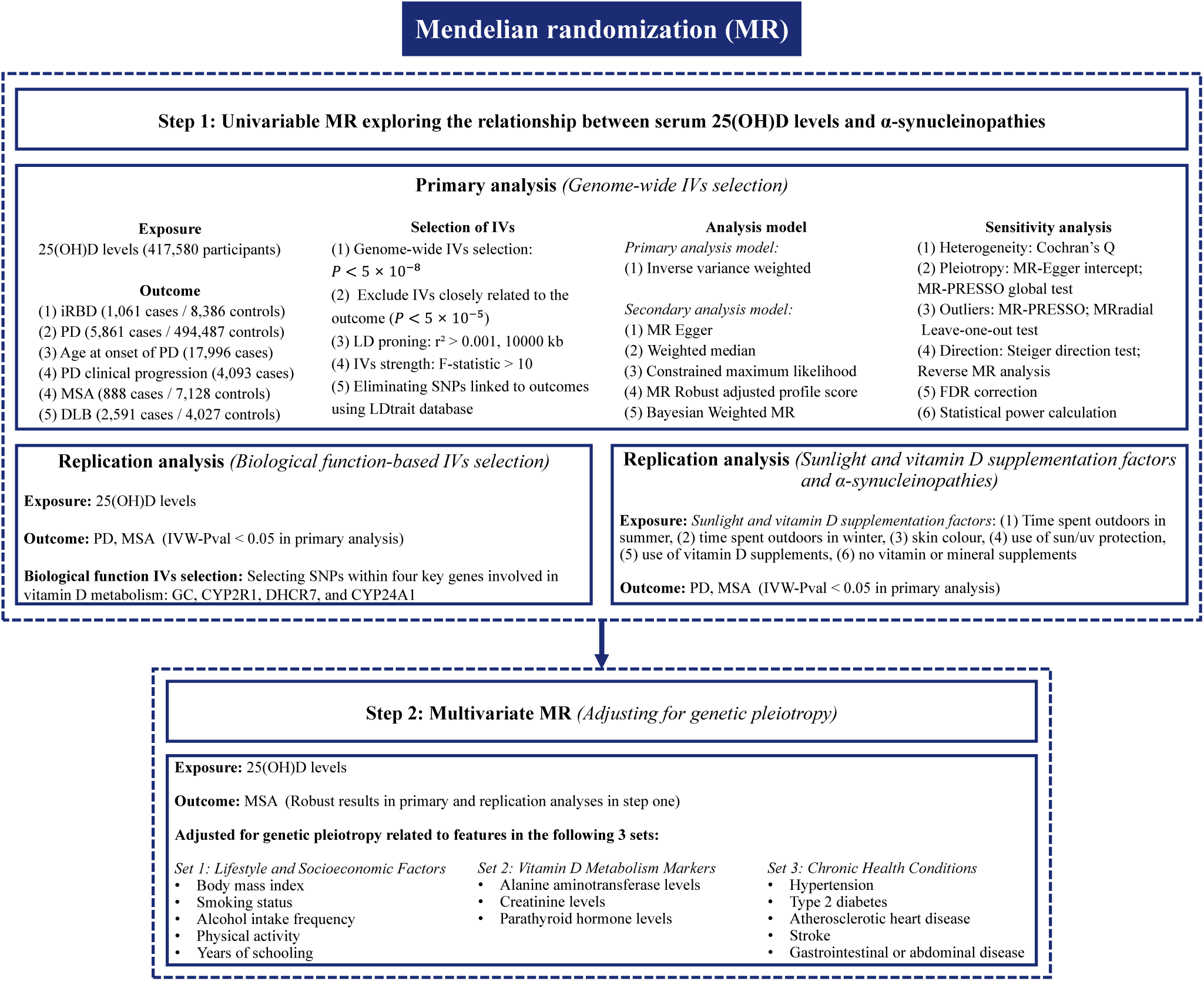

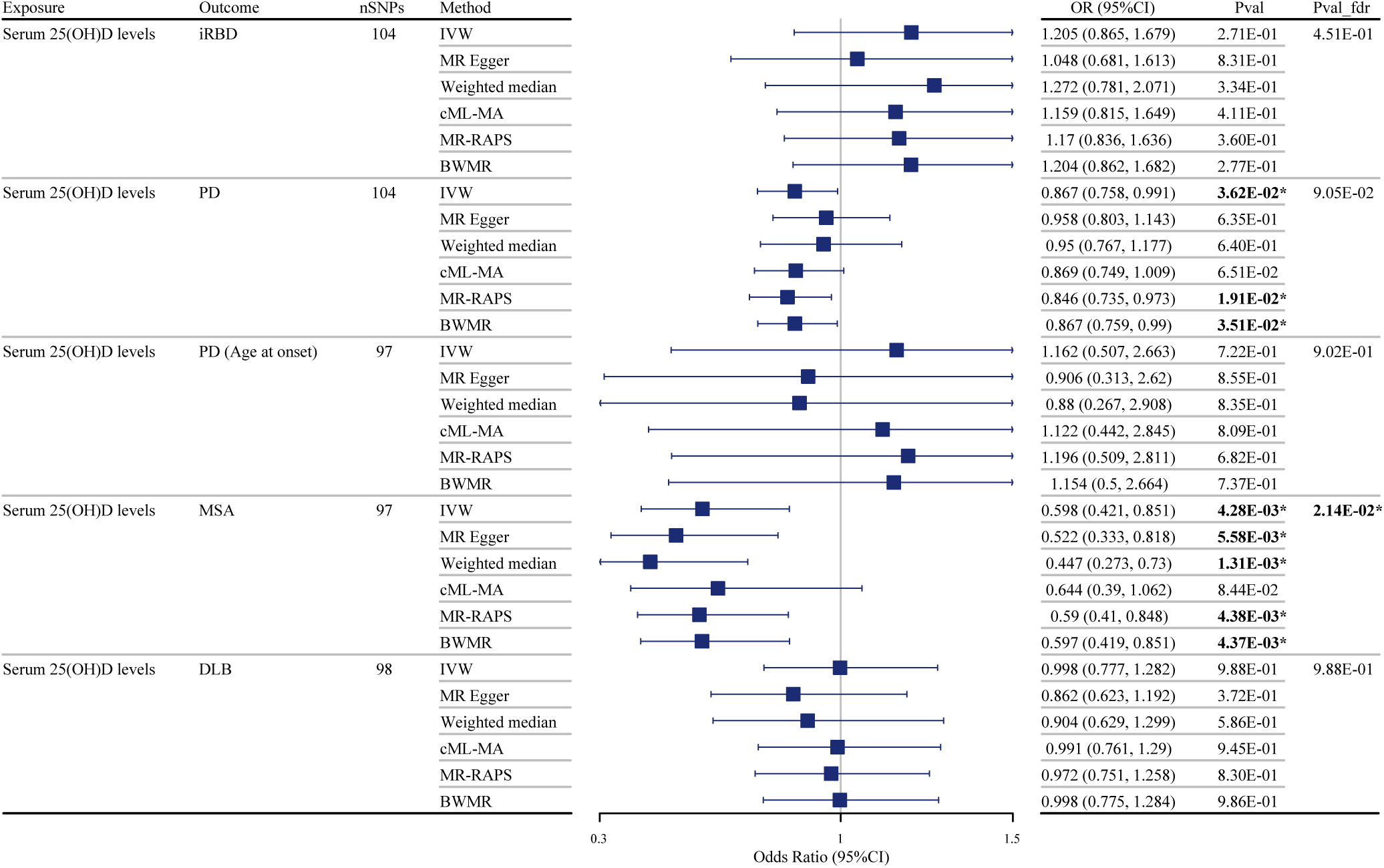

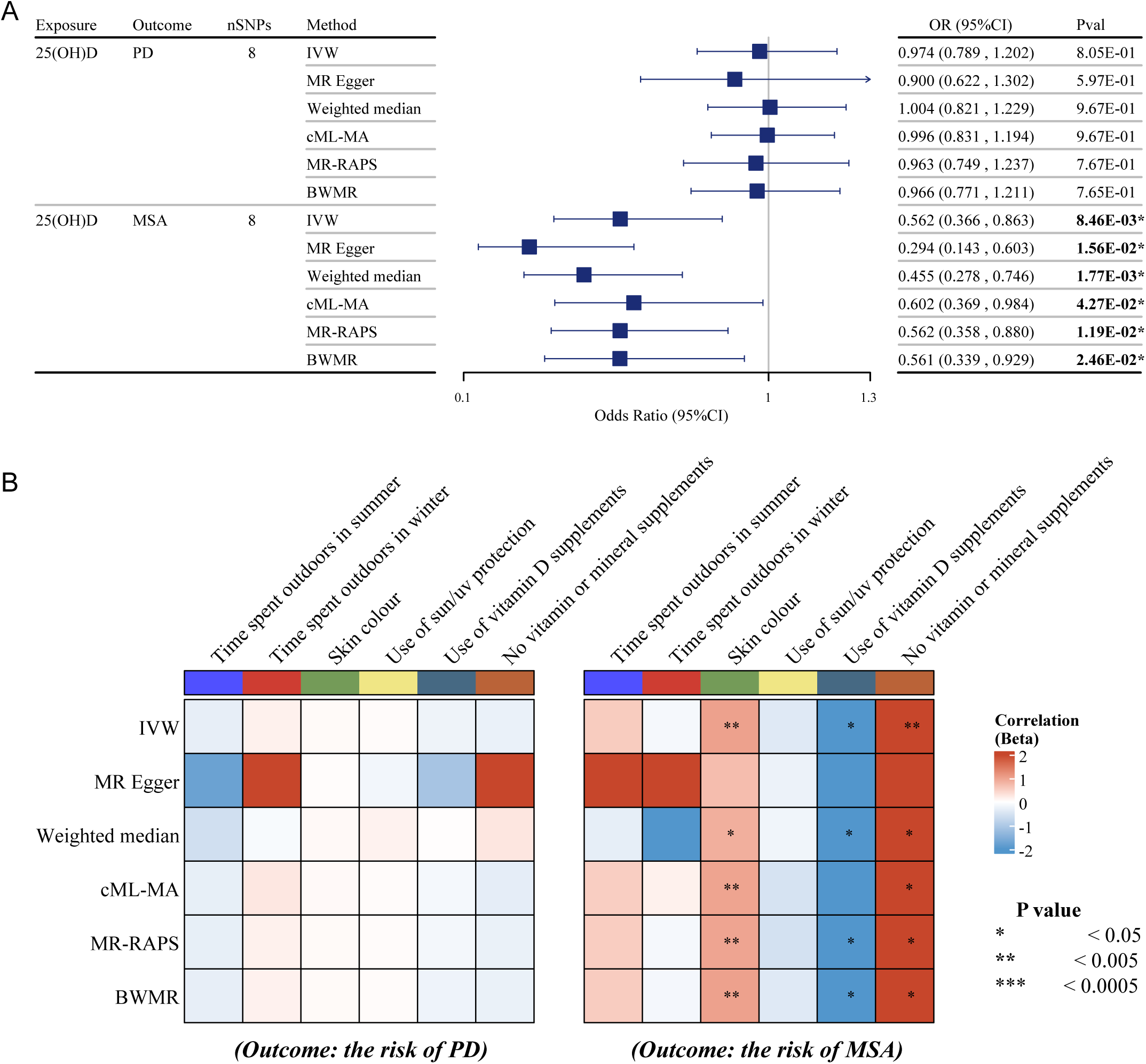

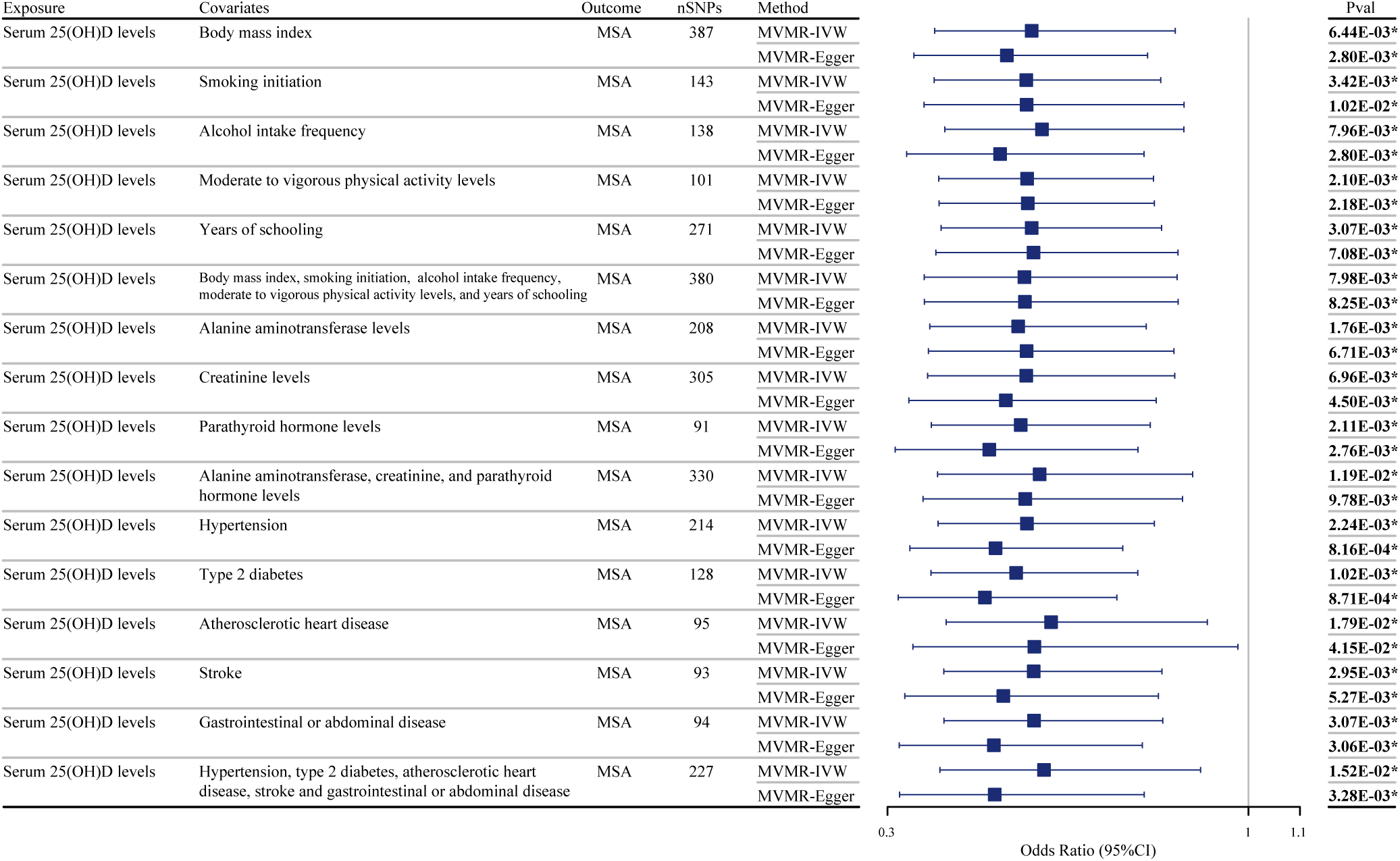

## Notes

### Competing Interest Statement

The authors have declared no competing interest.

### Author Declarations

The GWAS data sources used in this study are detailed in the Methods section and Supplementary materials - Table S1.

